# Defining indicators for disease burden, clinical outcomes, policies, and barriers to health services for migrant populations in the Middle East and North African region: a suite of systematic reviews

**DOI:** 10.1101/2023.07.11.23292496

**Authors:** Farah Seedat, Stella Evangelidou, Moudrick Abdellatifi, Oumnia Bouaddi, Alba Cuxart-Graell, Hassan Edries, Eman Elafef, Taha Maatoug, Anissa Ouahchi, Liv Mathilde Pampiri, Anna Deal, Sara Arias, Bouchra Assarag, Kenza Hassouni, Aasmaa Chaoui, Wafa Chemao-Elfihri, Mahmoud Hilali, Azeddine Ibrahimi, Mohamed Khalis, Mansour Wejdene, Ali Mtiraoui, Kolitha Wickramage, Dominik Zenner, Ana Requena-Mendez, Sally Hargreaves, the MENA Migrant Health Working Group

## Abstract

**Introduction:** The Middle East and North Africa (MENA) is characterised by high and complex migration flows, yet little is known about the health of migrant populations, their levels of under-immunisation, and access to healthcare provision. Data are needed to support regional elimination and control targets for key diseases and the design and delivery of programmes to improve health outcomes in these groups. This protocol describes a suite of seven systematic reviews that aim to identify, appraise, and synthesise the available evidence on burden and clinical indicators, policies, and barriers related to these mobile populations in the region.

**Methods and analysis:** Seven systematic reviews will cover two questions to explore: 1) the burden and clinical outcomes and 2) the policies, uptake, and the barriers for the following seven disease areas in migrants in the MENA region: tuberculosis, HIV and hepatitis B and C, malaria and neglected tropical diseases, diabetes, mental health, maternal and neonatal health, and vaccine-preventable diseases. We will search electronic databases for studies in any language (year 2000 to 2023), reference-check relevant publications, and cross-check included studies with experts. We will search for grey literature by hand searching key databases and websites (including regional organisations and MoH websites) for country-specific guidelines and talking to our network of experts for local and regional reports and key datasets. We will assess the studies and policies for their quality using appropriate tools. We will meta-analyse the data if they are of sufficient volume and similarity by disease outcome. Where meta-analysis is not possible and where data are on policy, we will narratively synthesise the evidence using summary tables, figures, and text.

**Ethics and dissemination:** We anticipate disseminating the findings through peer-reviewed publications, conferences, and other formats relevant to all stakeholders. We are following PRISMA guidelines and protocols will be registered on PROSPERO.

**STRENGTHS AND LIMITATIONS OF THE STUDY:**

- Explicit, transparent, and recommended systematic review methodology.
- Comprehensive search strategy with no language restrictions and extensive grey literature searching online and through expert government and non-government support.
- International team with multidisciplinary expertise on migrant health, diseases, policymaking, and methodology.
- Data prior to the year 2000 will be excluded.
- Meta-analysis may not be possible due to heterogeneity, and we anticipate that most of the data will come from grey literature.

## INTRODUCTION

The Middle East and North Africa (MENA) is marked by political and economic instability, extended conflict, and high and complex migrant flows. Several countries in this region are points of origin, transit, and destination for a varied group of migrants. More than 40 million migrants reside in the region,^1^ including 12.6 million internally displaced people (IDPs), 2.4 million refugees, 251,800 asylum seekers and 370,300 stateless persons at the end of 2022.^2^ The current conflict in Sudan has intensified the situation, with reports of over 1.4 million people becoming newly displaced (approximately 476,811 fleeing to neighbouring countries).^3^

Migrants are a heterogeneous group; while some are resilient throughout the migration cycle, have sufficient access to services in host countries and good health outcomes, others in the region live and work in precarious conditions.^4^ Some may face individual and system level barriers in accessing healthcare.^5 6^ Efforts have been made to enable access to affordable, acceptable, culturally sensitive, and good quality healthcare for migrants, and include them in national health programmes. For example, Morocco has multisectoral programmes through a national immigration and asylum strategy since 2013 to improve the health and well-being of all migrants, which includes the right of access to free or low-cost essential healthcare under the same conditions as Moroccans.^7 8^ However, migrants may still face economic barriers such as the cost of medication and complimentary tests, which may not be covered, as well as language and cultural barriers, a fear of deportation, racism, and discrimination.^5 9^ Non-governmental organisations and United Nations (UN) agencies often attempt to close the gap in healthcare provision, but these services often remain insufficient.^6 10^ This context can leave some migrant groups disproportionately affected by various health conditions, with poor morbidity and mortality outcomes.^11^

Globally, it is reported that certain migrant categories may encounter infectious diseases such as tuberculosis (TB) and HIV/AIDS along their journey and in their host countries, and they may be at risk of malaria and neglected tropical diseases for which health professionals may be unfamiliar. There is also evidence that they are disproportionately affected by the burden and consequences of COVID-19.^12^ Likewise, some migrants, especially refugees and asylum seekers, can be at risk of non-communicable diseases (NCDs), which are often diagnosed late and can be causes of premature mortality.^6 11 12^ In addition, depending on their lived experiences during the migratory trajectory and the contextual factors in the host country, some migrants may suffer adverse mental health outcomes.^12^ Refugee and migrant women tend to have less access to maternal and child health services and are at a higher risk of negative outcomes during pregnancy and delivery than women in host populations.^12^ Migrants are also considered to be an under-immunised group, missing vaccines, doses, and boosters, as children and adolescents because of their mobility, with WHO calling for greater emphasis to be placed on vaccination across the life-course in marginalised groups.^13 14^

There is a paucity of data mapping the burden of diseases and under-immunisation in migrants in the MENA,^12^ however, some small studies in some countries suggest that migrants may be at higher risk for some diseases. For example, in Lebanon, migrants are considered a vulnerable group for HIV as the political and economic situation has led to an increase in high-risk behaviours,^15^ while cross-border mobile populations in the ports of the Red Sea and adjacent transport corridors are found to be at a higher risk of HIV transmission in the Horn of Africa and Arabian Peninsula.^16^ Similarly, high rates of hepatitis B and C have been found in newly arriving sub-Saharan migrants in Libya (23.4% and 31.2% respectively),^17^ and migration has been identified as partly explaining the slowing of the decline in TB and the re-introduction of malaria in the region,^18^ as well as outbreaks of neglected tropical diseases such as leishmaniasis and cysticercosis/taeniasis in Lebanon.^19 20^

The evidence on NCDs is similar. A scoping review in 2019 on Syrian refugees found only two studies that were investigating prevalence – one in Lebanon and one in Jordan.^21^ The studies found that almost one in two households had a member with an NCD; the two most common NCDs were hypertension and Type 2 diabetes mellitus. In Lebanon, the proportion of NCDs was higher in the refugees than the host community (60.2% vs 50.4%, respectively). A retrospective study in Qatar similarly found that diabetes and hypertension were higher in migrants who had just arrived (<6 months) compared with longer durations.^22^ There is also some evidence of a high burden of psychiatric disorders, such as generalised anxiety disorder and post-traumatic stress disorder in Lebanon, Sudan, and Egypt among refugees and IDPs with pre-migration exposure to armed conflict.^23^ The Gulf Cooperation Council reported high rates of psychosis and suicide among migrant domestic workers coming from Indian Subcontinent and South East Asia.^24^ With respect to maternal and neonatal conditions, a study in Lebanon found that the odds of very preterm birth and other serious antenatal complications were higher for most migrant women compared to host women.^25^

There is equally little information on the policies and interventions (e.g., screening, diagnostic, treatment, or vaccination services) available to migrants in the MENA. Indeed, when exploring the NCD policies for urban refugees, a scoping review in 2020 concluded that there is a scarcity of research on national policy, the prevention of NCDs, and the perspective of refugees.^26^ Likewise, there is little comprehensive data about infectious disease policies among migrants in the MENA. Two Sudanese studies reported a low vaccination coverage among migrant and internally displaced children against major vaccine preventable diseases (VPDs).^27 28^ In countries such as Egypt, Iraq, Jordan, Lebanon, Morocco and Tunisia, migrants are included in vaccination policy irrespective of their residency status. However, in some countries, some groups of migrants often cannot access sufficient vaccinations, and NGOs are left to fill this gap, especially for irregular migrants.^13^

The lack of comprehensive data on the burden of diseases among migrants in the MENA makes it difficult to respond to the unmet healthcare needs of the migrant population.^1 29^ Likewise, the extent to which migrants are included in national policies and the barriers they may face could undermine the goals to achieve the legal and human right of universal health coverage, ensure health security for all, and the eliminate some of these diseases. Efforts are underway to make integrated migrant health information systems a reality in the MENA. Since 2015, the International Organization for Migration (IOM) has steered projects fostering health and protection to vulnerable migrants transiting through Morocco, Tunisia, Egypt, Libya, Yemen and more recently, Sudan.^30^ The current phase aims to steer the development of a migrant health country profile tool (MHCP-t) for the region, an innovative digital mechanism to source country level data on health indicators, health policies, and healthcare access across multiple communicable and non-communicable disease areas. The MHCP-t will enable a systematic assessment of disease impact on migrants and identify country-level hotspots and gaps in prevention efforts by integrating information from routine health data, registries, surveillance programmes, humanitarian stakeholders, and civil society on health aspects of migrant populations (see figure 1).

**Figure 1.**
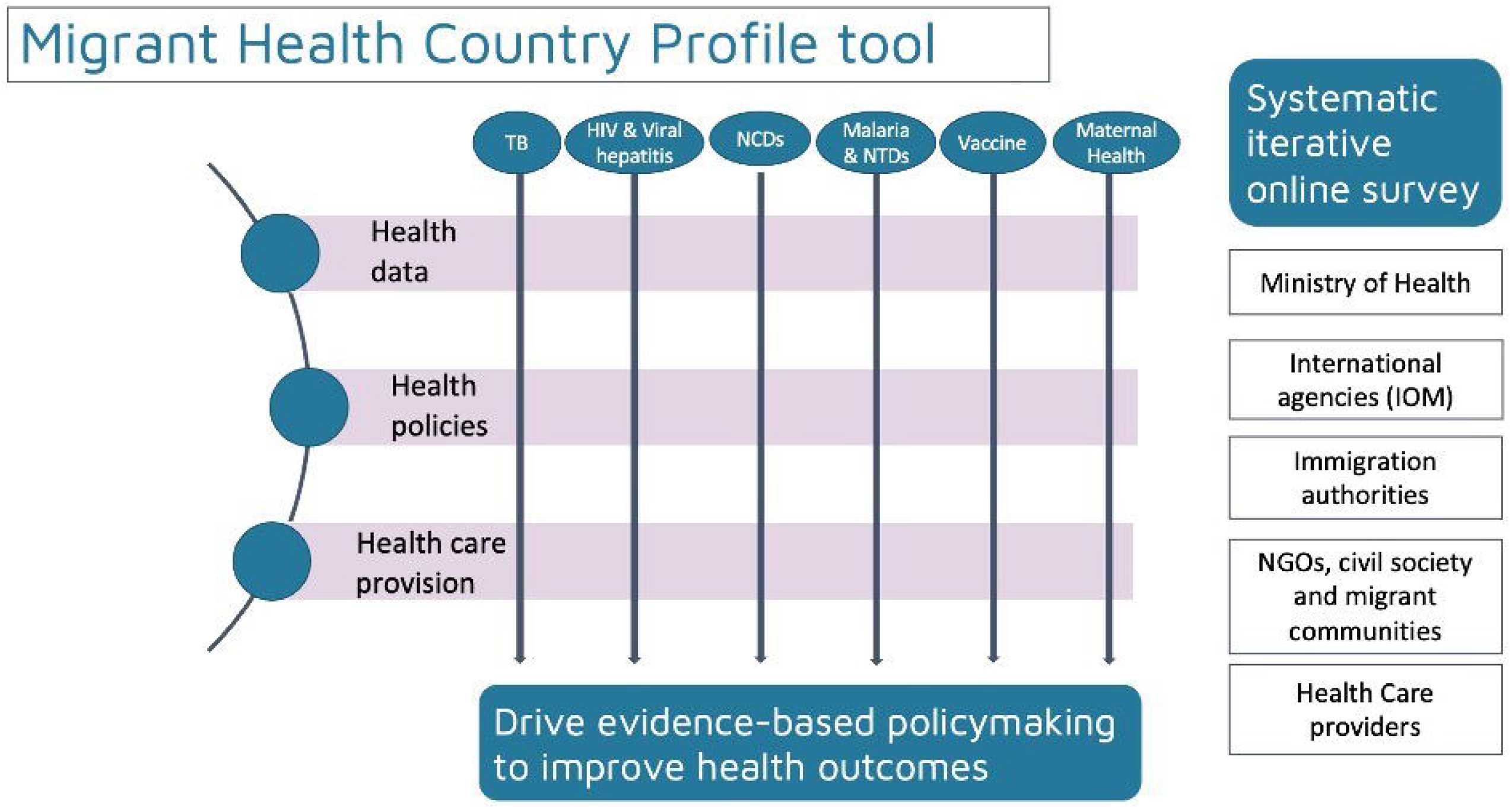
Schematic illustration of the Migrant Health Country Profile Tool (MHCP-t)

A preliminary step in the development of the MHCP-t is to map out the state of the evidence on migrant health data and policies in the MENA and to inform the development of key indicators for disease burden and clinical outcomes, policies, and access for all relevant disease areas of the MHCP-t. To date, there have been no attempts to systematically synthesise the disease, policy, or access indicators in the literature about key diseases among migrant populations in the MENA region. This suite of systematic reviews will aim to systematically identify, appraise, and synthesise the empirical evidence on key diseases among migrant populations in the MENA.

### Research questions and objectives

There are two overarching research questions that will be addressed in seven systematic reviews on seven disease areas to comprehensively map the state of the evidence of migrant health in the MENA: TB (1), HIV and hepatitis B and C (2), malaria and neglected tropical diseases (3), diabetes (4), mental health (5), maternal and neonatal health (6), and VPDs (7).

1. What data are available on the disease indicators related to each of the seven disease areas in migrant populations in the MENA region? Objectives:
  a. Synthesise the burden of TB, HIV, hepatitis B and C, malaria, neglected tropical diseases, diabetes, mental health, maternal and neonatal health conditions, and VPDs in migrant populations in the MENA region.
  b. Synthesise the (intermediate and final) clinical outcomes of TB, HIV, hepatitis B and C, malaria, neglected tropical diseases, diabetes, mental health, maternal and neonatal health conditions, and VPDs in migrant populations in the MENA region.
  c. Examine the quality of evidence on the burden, clinical process, and final health outcomes of TB, HIV, hepatitis B and C, malaria, neglected tropical diseases, diabetes, mental health, maternal and neonatal health conditions, and VPDs in migrant populations in the MENA region.
2. What is the policy response for each disease area related to migrant populations in the MENA region, what is the uptake, and what are the barriers in accessing these policies? Objectives:
  a. Synthesise the prevention and/or treatment policies for TB, HIV, hepatitis B and C, malaria, neglected tropical diseases, diabetes, mental health, maternal and neonatal health conditions, and VPDs in migrant populations in the MENA region.
  b. Synthesise the uptake of prevention and/or treatment services for TB, HIV, hepatitis B and C, malaria, neglected tropical diseases, diabetes, mental health, maternal and neonatal health conditions, and VPDs in migrant populations in the MENA region.
  c. Synthesise the evidence on the barriers for accessing prevention and/or treatment services for TB, HIV, hepatitis B and C, malaria, neglected tropical diseases, diabetes, mental health, maternal and neonatal health conditions, and VPDs in migrant populations in the MENA region.
  d. Examine the quality of the prevention and/or treatment policies for TB, HIV, hepatitis B and C, malaria, neglected tropical diseases, diabetes, mental health, maternal and neonatal health conditions, and VPDs in migrant populations in the MENA and cross-compare between countries in the MENA region.
  e. Examine the quality of evidence on the uptake and barriers of prevention and/or treatment policies for TB, HIV, hepatitis B and C, malaria, neglected tropical diseases, diabetes, mental health, maternal and neonatal health conditions, and VPDs in migrant populations in the MENA region.

We will disaggregate by, and investigate potential sources of heterogeneity for, country of origin, migrant type (i.e., labour, asylum seekers, refugee), age, and sex where feasible for all objectives, and cross-compare findings across countries in the MENA region.

**Panel 1.**
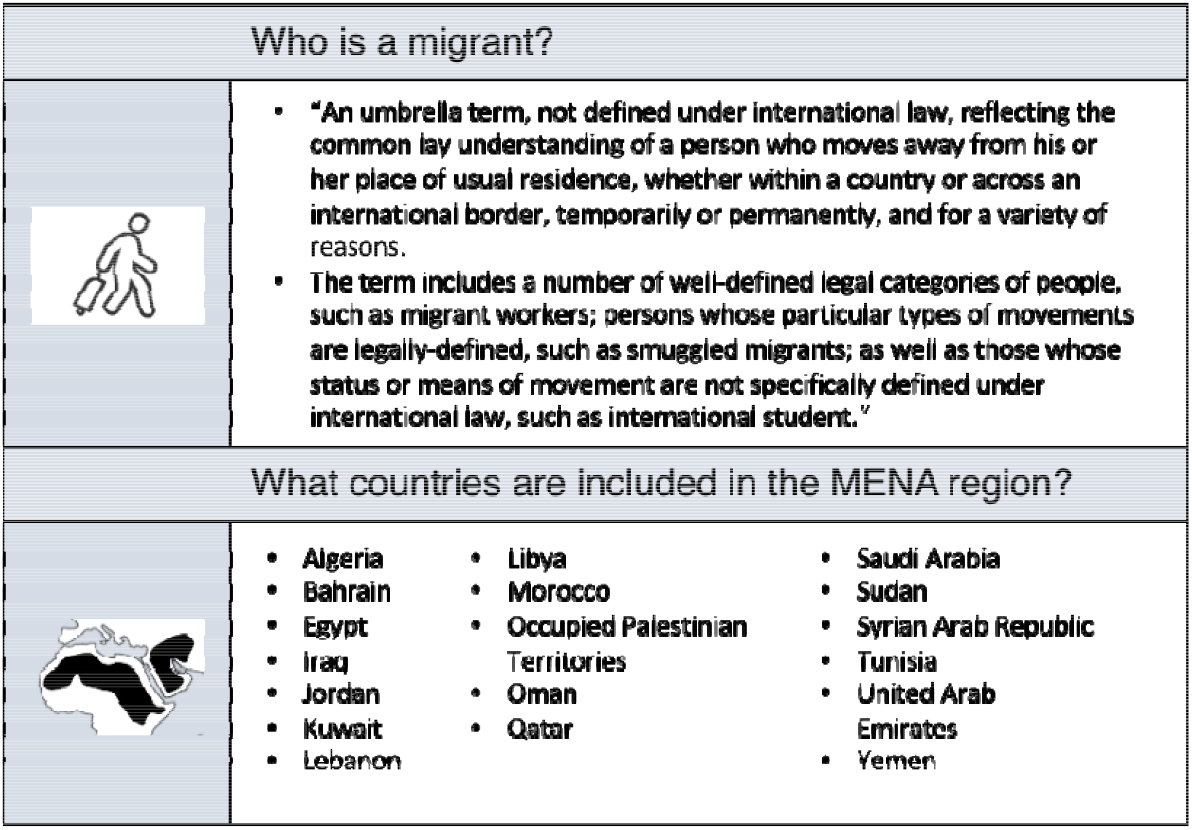
Definitions used in this project, adopted from the International Organization for Migration^31 32^.

## METHODS AND ANALYSIS

The seven systematic reviews will be reported according to recommendations from the Preferred Reporting Items for Systematic Review and Meta-analysis Protocols (PRISMA-P) 2020 statement.^33^ We will register the systematic reviews protocols at the International Prospective Register of Systematic Reviews (PROSPERO).

### Search strategies

#### Electronic database searches

We will search electronic databases to identify peer-reviewed literature. Scoping searches have been undertaken to inform the development of the search strategies. An iterative procedure was used, with input from all authors including an information scientist, recommended search filters, and previous reviews. The searches are separated for each disease area. They combine three sets of search terms using both free text words in title, abstract, or keyword heading word, and MeSH terms through boolean operators OR within each set and then AND to combine the sets. The first set is made up of search terms for migrants, the second set is made up of search terms for each disease area, and the third set is made up of search terms for the MENA region. The search strategies are limited to humans and restricted to the year 2000 as migratory flows and disease burden change over time, and we are interested in recent migration flows and recent policies. There is no restriction on language. A copy of the overarching search strategy used in the major databases is provided in Table 1, and the strategies specific to each disease area (i.e., terms for diseases) are presented in the supplementary material.

**Table 1.** Overarching search strategy for all reviews.

|  |  |  |
| --- | --- | --- |
| **1** | **Migrant** | (alien* or asile or asylum* or (border* adj2 cross*) or (countr* adj3 origin*) or diaspora or displace? or displacement* or emigrant* or emigration or expat? or expatriate? or foreigner* or foreign-born* or foreign background* or foreign population* or immigrant* or immigration or migrant* or migration or naturalized citizen* or new* arriv* or newcomer* or new-comer* or nomad* or non-citizen* or nonnative* or non-native* or nonnational or non-national or nonresident or non-resident* or resettlement* or re-settlement* or refugee* or settler* or squatter* or undocumented worker*).ti,ab,kf.<br>or exp Human Migration/ or exp "Emigrants and Immigrants"/ or "Transients and Migrants"/ or Refugees/ or Refugee Camps/ |
| **2** | **Disease** | Separate for each strategy (see supplementary material) |
| **3** | **Countries** | (Abu Dhabi or Ajman or Algeri* or Arab* or Bahrain* or Bahreini* or Dubai or Egypt* or Emirat* or Fujairah or Gaza* or Golf* or Gulf* or Ifriqiya* or Irak* or Iraq* or Jorda* or Jumhuuriya* or Koweit* or Kuwait* or Kuwayt* or Leban* or Liban* or Liby* or Lubnan* or Maghr* or Maroc* or Maser* or Masr or Misr or MENA or Middle East* or Morocc* or North* Afric* or Oman* or Palestin* or Qatar* or Saudi* or Sharjah or Soudan* or Sudan* or Syria* or Syuri* or Tunis* or Uman* or Umm Al-Quwain or West Bank or Yemen*).ti,ab,kf.Exp middle east/ Exp Africa, Northern/<br>exp Middle East / or exp Africa, Northern/ |
| **4** | **Combine for indicators** | 1 AND 2 AND 3 |
| **5** | **Limit to humans** | (animals not humans).sh.<br>4 not 5 |
|  | **Limit to 2000** | Limit 5 to yr="2000-Current" |

The bibliographic databases we will search are MEDLINE (Ovid); EMBASE (Embase.com); Web of Science (Clarivate), CINAHL (Ebsco), Index Medicus for the Eastern Mediterranean Region (globalindexmedicus.net), Qscience (qscience.com).

#### Grey literature searches

We will search various sources to identify literature that has not been published in peer-reviewed journals. Firstly, we will search the reference lists of the final list of included studies at full text as well as systematic reviews that meet our criteria that we have sourced from searches of the electronic databases. Secondly, we will search international and regional websites related to migration and health, such as the IOM, WHO, UNHCR, Red Cross and other relevant UN bodies, Médecins San Frontières (MSF) and other key regional NGOs, and national websites for each country in the MENA, particularly ministries of health for guidelines, policies, reports, and unpublished datasets. We will use snowballing methodology and search any relevant websites we find from these initial websites, as necessary. Once we have a full list of included papers and websites, we will share this with a panel of country-specific and regional experts to review and identify any further data sources that have not been captured.

### Study screening and selection

The eligibility criteria by disease area are summarised separately in the Table 2 for disease indicators (question 1) and Table 3 for policy and access (question 2). For disease indicators (question 1), we will include papers that are on the burden (e.g., prevalence or incidence) or intermediate (e.g., coverage or completion of interventions such as screening or treatment) or final (e.g. morbidity, mortality, quality of life) clinical outcomes for TB, HIV, hepatitis B and C, malaria, neglected tropical diseases, diabetes, mental health, maternal and neonatal health conditions, and VPDs in migrant populations in the MENA region. For the policy-related data (question 2), we will include papers that contain a description of the policies themselves, uptake of the health services mentioned in the policies and determinants of any under-usage, and facilitators or barriers in accessing the health services mentioned in the policies for the diseases in migrant populations in the MENA region. Definitions for migrant and the MENA region are described in panel 1.

**Table 2.**
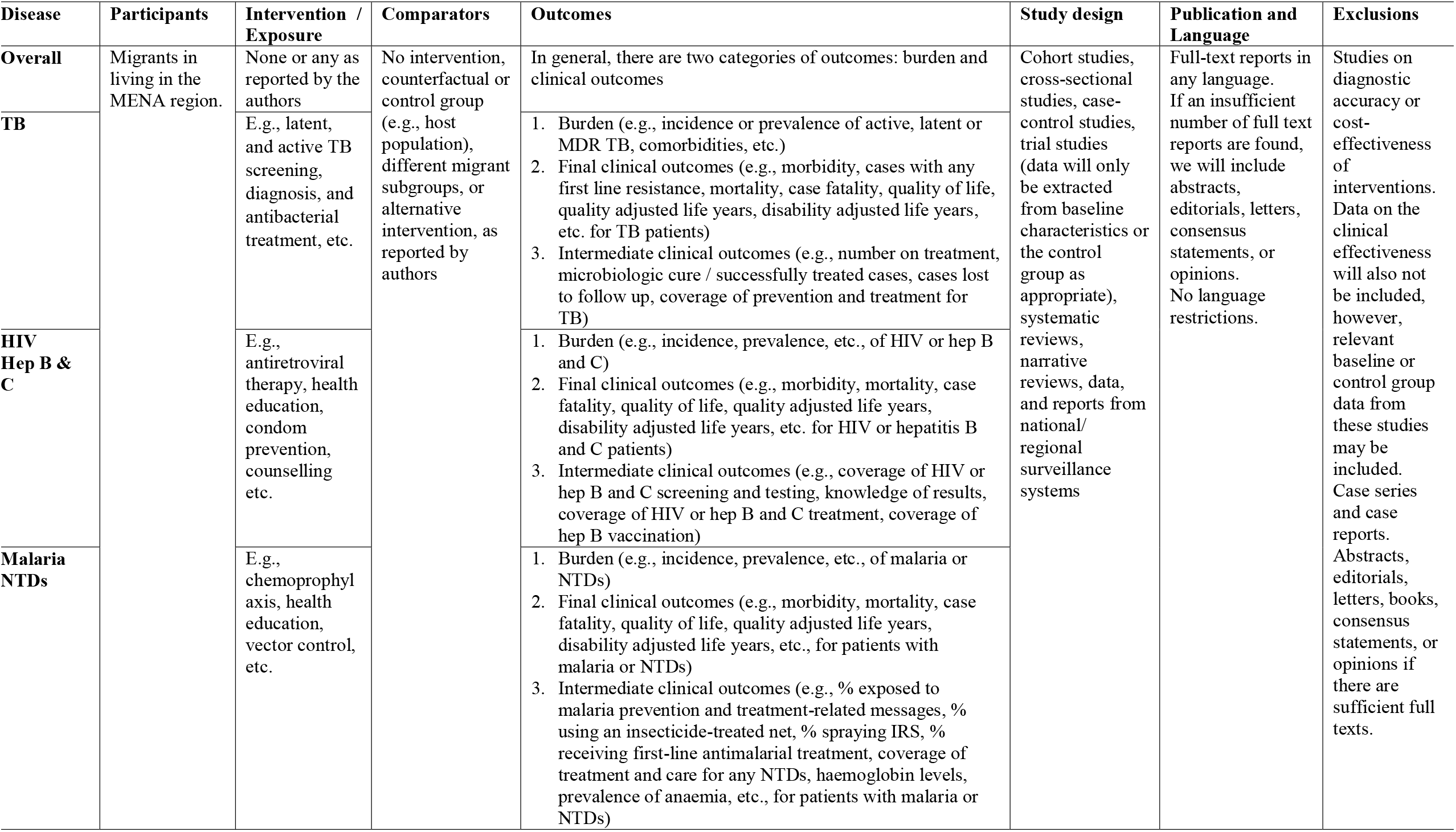
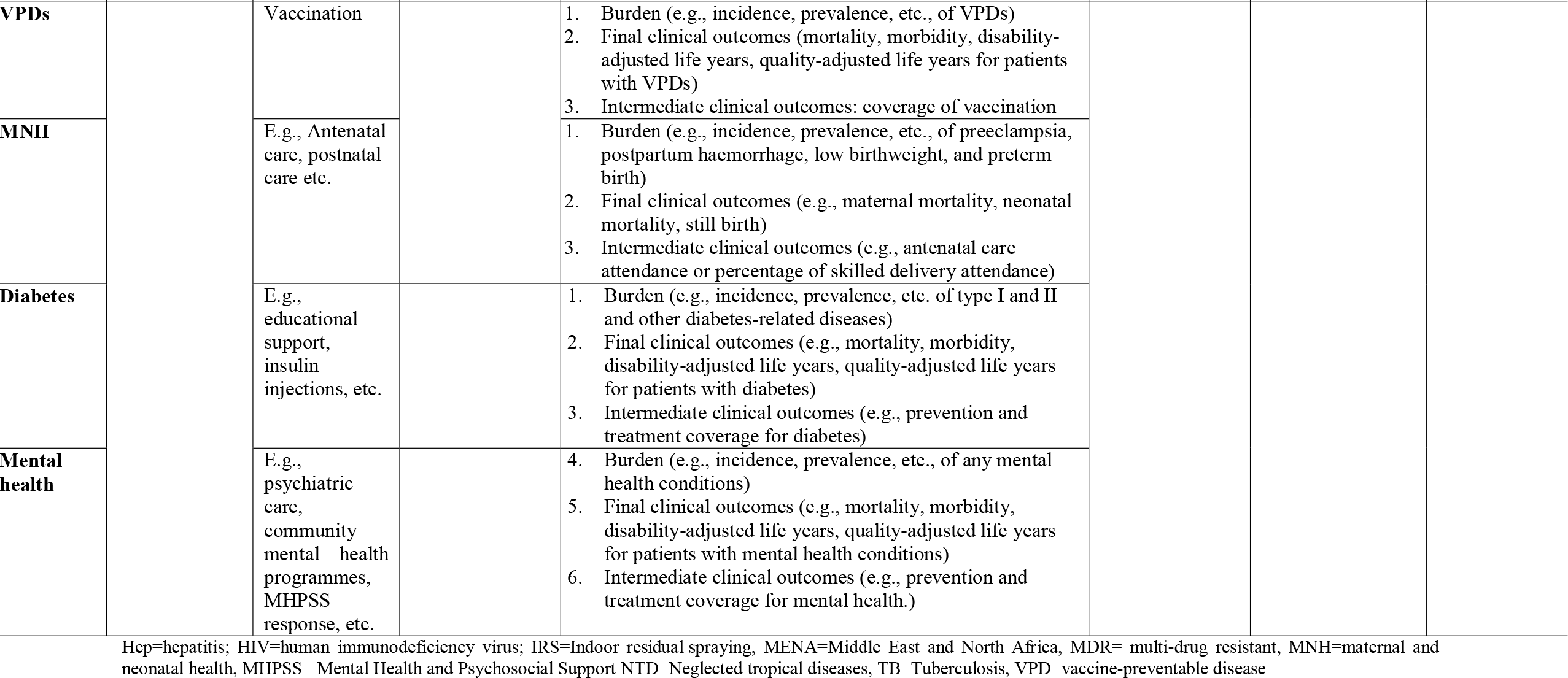
Study eligibility criteria for disease indicators (question 1)

**Table 3.**
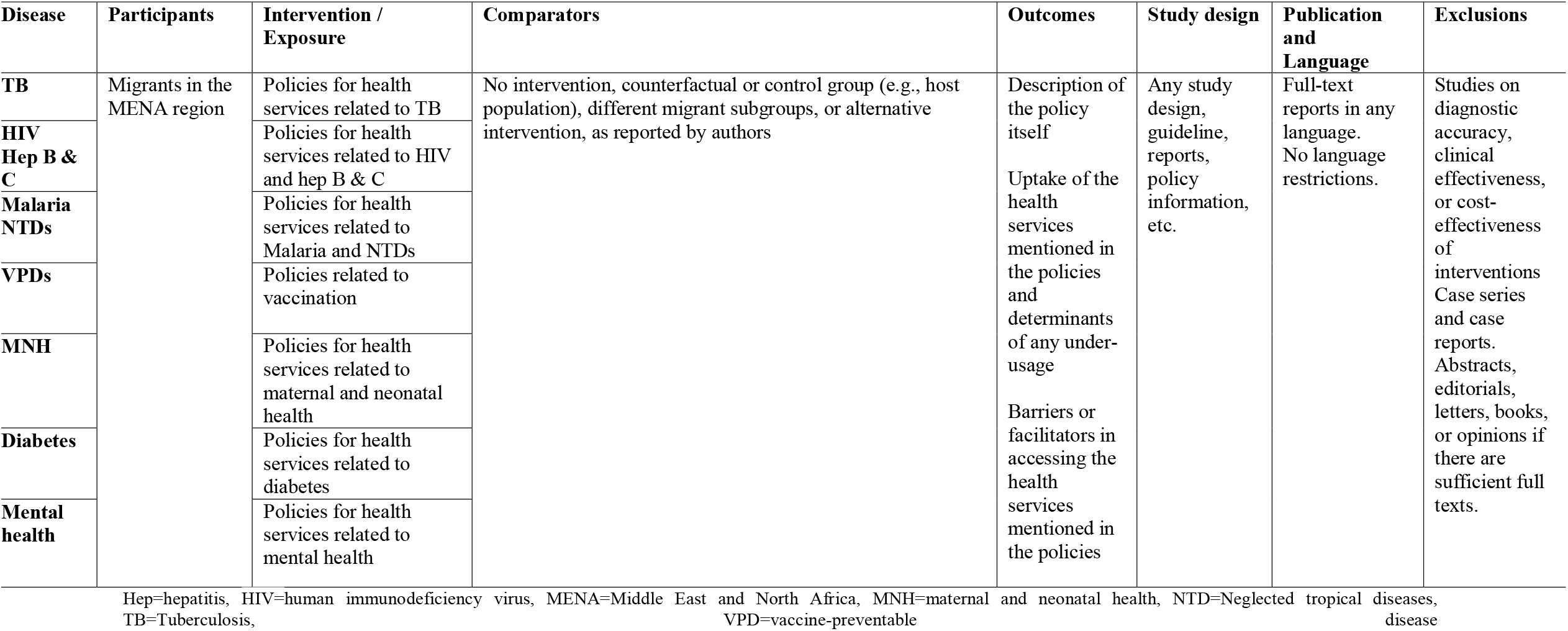
Study eligibility criteria for policy indicators (question 2)

For disease areas that concern multiple diseases, we have prioritised those that are of greater concern in the MENA region. We are including the following vaccine preventable diseases: cholera, COVID-19, dengue, diphtheria, hepatitis A, hepatitis B; haemophilus influenza type b (Hib), human papillomavirus (HPV), influenza, measles, meningococcal diseases, mumps, pertussis, pneumococcal disease, poliomyelitis, rabies, rubella, rotavirus, tetanus, tuberculosis, typhoid, and varicella. For neglected tropical diseases, we will include buruli ulcer, dengue and chikungunya, dracunculiasis (Guinea-worm disease), echinococcosis, human African trypanosomiasis (sleeping sickness), leishmaniasis, leprosy (Hansen’s disease), lymphatic filariasis, mycetoma, and other deep mycoses, onchocerciasis (river blindness), rabies, scabies and other ectoparasitoses, schistosomiasis, soil-transmitted helminthiases, snakebite envenoming, taeniasis/cysticercosis, trachoma, and yaws and other endemic treponematose. We are including any mental health condition, and for NCDs, we have chosen diabetes.

With respect to interventions and comparators, for disease indicators, we will include studies in which any intervention or exposure is reported, or studies where no intervention is reported if the study is about prevalence or incidence alone. For policy-related data, we will include any health interventions relevant for the disease area. For both the systematic and the policy reviews, we will include studies that have no comparator or any type of comparator (i.e., a counterfactual group, control group, reference group for an exposure, different migrant subgroup, or an alternative intervention). For publication type, we will include full texts of observational studies, data from national and regional surveillance systems, and trials for the systematic review on indicators, although for trials, we will only extract data from the control group or from baseline characteristics, as appropriate. We will include systematic reviews and narrative reviews to search for information and policies from these papers. For policy-related data, we will include full text papers from any study design and guidelines and policy information. We will exclude studies on diagnostic accuracy, cost effectiveness of interventions, case series and case reports, and exclude abstracts, editorials, books, or opinions, if there are a sufficient number of other publications. We will have no language exclusions.

Identified references will be downloaded to bibliographic management software, Rayyan, and de-duplicated. Two reviewers will independently screen the titles and abstracts of all identified records (screening level I). Full text reports of all potentially relevant records identified at screening level I will be obtained and assessed independently by two reviewers using the same study eligibility criteria (screening level II). Any disagreements over inclusion/exclusion at screening level I and II will be resolved by discussion between the two reviewers, with the involvement of a third reviewer if necessary. We will document the study flow and reasons for exclusion of full text papers in a PRISMA study flow diagram (see supplementary material).

### Data extraction

Two reviewers will independently extract relevant data using an a priori defined extraction sheet that will be piloted and refined before implementation. Data extracted will be cross-checked and any disagreements will be resolved by discussion, with the involvement of a third reviewer if necessary.

Data extracted for the systematic review on indicators (question 1) will include study (e.g., author, country, publication year, design, setting, sample size, follow-up duration), participant (e.g., type of migrant and definition used, country of origin, time in the country, socio-demographic characteristics, living situation [camp setting, community], study eligibility criteria), health or disease indicators including their definitions (e.g. description of the intervention if it is about testing or education or treatment), the outcome measures (e.g. frequency, percentages, 95% CIs, etc.), results reported for each outcome, and any adjustment conducted for confounding of the outcomes. From each study that has adjusted and unadjusted analyses, we will prioritise the adjusted analysis if data allow. Any missing statistical parameters of importance and variability measures (e.g., 95% CIs) will be estimated, if data permit. All calculated or derived data will be denoted as ‘calculated’ and will be incorporated in the extraction sheets.

Data extracted for the systematic review on policies (question 2) will include study (e.g., author, country, publication year, design, setting, sample size, follow-up duration), participants included in studies or population covered by policy (e.g., type of migrant and definition used, socio-demographic characteristics, living situation [camp, setting, community] study eligibility criteria), summary of interventions (in the study or covered by policy) and comparators (for studies on uptake and barriers), outcome (e.g. policy, uptake, and/or barriers), results reported for each outcome (e.g. summary of policy, level of authority [national, regional, local], implementation [legal, recommendation, guideline] for policy outcome, list of barriers for barriers outcome, and frequency, percentages, and 95% confidence intervals for uptake outcome). Any missing statistical parameters of importance and variability measures (e.g., 95% CIs) will be estimated, if data permit. All calculated or derived data will be denoted as ‘calculated’ and will be incorporated in the extraction sheets.

### Risk of bias

Two independent authors will appraise the risk of bias for each included paper. For peer-reviewed literature, we will use the appropriate Joanna Briggs Institute (JBI) tool for each study design and for grey literature records, we will use the AACODS Checklist.^34 35^ We will assess the quality of the policies using the most appropriate AGREE tool – AGREE II or AGREE HS.^36^ The quality appraisals will be cross-checked and any disagreements will be resolved by a consensus-based discussion, with the involvement of a third reviewer if necessary. The individual item-specific quality assessment ratings for each study will be tabulated. Records will not be excluded based on quality assessment, but the appraisal will contribute to the synthesis and the discussion.

### Data synthesis and analysis

#### Disease indicators (question 1)

If the studies are sufficiently similar, we will combine the data using a proportional meta-analysis via a random effects model due to the anticipated heterogeneity that may result from the differences in methodology and study settings.^37^ There will be a separate pooled estimate for each disease indicator / outcome. If there are sufficient data, there will be a separate pooled estimate of studies with adjustment and studies without adjustment. If we do not have sufficient data, adjusted and unadjusted studies will be pooled together for each disease indicator / outcome.

We will assess heterogeneity among studies by inspecting the forest plots and using the chi-squared test for heterogeneity with a 10% level of statistical significance and using the *I*^*2*^ statistic where we interpret a value of 50% as representing moderate heterogeneity. We will assess the possibility of publication bias by evaluating funnel plot asymmetry and will also be conducted using an adjusted Egger’s regression asymmetry test as a formal statistical test for publication bias for outcomes with ten or more studies. We will perform a leave-one-study-out sensitivity analysis to determine the stability of the results. This analysis will evaluate the influence of individual studies by estimating the pooled analyses in the absence of each study.

If there are sufficient data, we will investigate potential sources of heterogeneity, using meta-regression, and incorporating the following covariates in each model: type of migrant (labour, asylum seeker, refugee, undocumented, etc.); setting/housing (camps, community, detention etc.); comorbidities; country of birth; age and sex. To assure confidence in the results of the meta-analyses, if there are sufficient studies, we will include the following sensitivity analyses: only include studies rated low risk of bias, only include studies that had adjustment for confounding factors, and only include prospective studies.

When studies cannot be combined for meta-analysis due to significant clinical heterogeneity, such as differences in participant characteristics, outcome measurements, etc., narrative syntheses will be conducted and results of individual studies will be displayed in tables, texts, and figures as appropriate, to enable a succinct summary of evidence.

#### Policy-related information (question 2)

For uptake, if the studies are sufficiently similar, we will combine the data exactly as specified above for indicators. For policies, narrative syntheses will be conducted, and results of individual studies will be displayed in tables, texts, and figures as appropriate, to enable a succinct summary of evidence. Likewise, if studies on uptake cannot be combined by meta-analysis due to substanial heterogeneity, such as differences in participant characteristics, interventions, etc., narrative syntheses will be used. We will use thematic analysis to group the facilitators and barriers reported across studies into themes and display the results in tables, texts, and figures, as appropriate.

All statistical analyses will be conducted in R statistical software (version 4.2.2).

## Data Availability

All data produced are available online in electronic databases or websites.

## ETHICS AND DISSEMINATION

There are no ethical or safety issues. The seven systematic reviews will identify and summarise the relevant evidence on the data on disease burden, clinical outcomes, policies, and barriers to access in migrant populations in the MENA region. The findings of the systematic reviews will be summarised along with the methodological quality of the studies. Strengths and limitations of the review will be discussed and gaps in the evidence will be highlighted. The findings of these seven reviews, and those of other similar reviews or reports (if identified), will be compared.

We aim to publish each of the individual systematic reviews in peer-reviewed journals as the findings have global relevance so a peer-reviewed journal will give us this reach. In addition, we will present these findings in oral and poster presentations in relevant conferences nationally in the MENA region and internationally. We also intend to report the findings to ministries of health in Morocco, Tunisia, and Sudan where we will be conducting the qualitative studies to continue the development of the MHCP-t. In addition, we will report some of these findings on our website for this project: the MENA Migrant Health project (http://www.menamigranthealth.org/). We will explore innovative ways to do public engagement work to disseminate the findings of this work to community organisations, migrant groups, NGOs, and others through our growing networks in the MENA region. We envisage that they may use these findings for advocacy work and to lobby for improved service provision locally and country wide.

The data on disease burden, clinical outcomes, policies, and barriers to access related to migrant health in the MENA region have not been systematically reviewed, yet they have important implications for the health and well-being of migrants and the health of the local populations. These findings will be discussed with a view to better inform the understanding of data in the MENA region and the indicators we should use in the MHCP tool. The findings will also contribute more widely as a basis for future research on migrant health in the region.

## ACKNOWLEDGEMENTS

We would like to thank Sabina Gillsund and Narcisa Hannerz for developing the search strategies and all the members of the Middle East and North Africa Migrant Health Working Group: Asad Adam (University of Gezira, Sudan), Adnene Ben Haj Aissa (Office National de la Famille et de la Population, Tunisia), Charles Agyemang (University of Amsterdam, Netherlands), Salma Altyib (Ministry of Health, Sudan), Ali Ardalan (WHO Regional Office for the Eastern Mediterranean), Ibrahim Bani (Ajman University, United Arab Emirates), Nuria Casamitjana (University of Barcelona, Spain), Luciana Ceretti (IoM MENA), Mohamed Douagi (Office National de la Famille et de la Population, Tunisia), Algdail Elnil (Sudan Organization Network for Peace & Development, Sudan), Gonzalo Fanjul (Institut for Global Health Barcelona, Spain), Fouad M Fouad (American University of Beirut, Lebanon), Chrifi Hassan (Ecole Nationale de Santé Publique, Morocco), Chiaki Ito (IoM MENA), Davide Olchini (Médecins du Monde MENA), Tarik Oufkir (Maroc Solidarité Médico-Sociale MS2, Morocco), Wafa Saidi (Ministry of Health, Tunisia), Sandra Santafé (Institut for Global Health Barcelona, Spain), Fatma Temimi (Office National de la Famille et de la Population, Tunisia).

## AUTHORS’ CONTRIBUTION

ARM and SH conceptualised the idea for this project. FS planned the overall methodology for the reviews, is coordinating the reviews, and led the write up of this manuscript. MA, OB, HE, EE, TM, and AO adapted the overarching methodology for each disease area and will be the first and second reviewers for the systematic reviews. AD and SAL contributed to the review on vaccination. KA, WA-E, MH, AI, MK, WM, AM, DZ, ARM, and SH are supervising the reviews. All authors were involved in the drafting and reviewing of the manuscript.

## PATIENT AND PUBLIC INVOLVEMENT

Members of the MENA Migrant Health Working Group, including clinicians and policymakers from the Ministries of Health, IOM, WHO, Médecins du Monde, and Maroc Solidarité Médico-Sociale MS2, have been involved in the design of this protocol.

