## Supplementary Material for "Defining indicators for disease burden, clinical outcomes, policies, and barriers to health services for migrant populations in the Middle East and North African region: a suite of systematic reviews"

### **Draft search strategies for the systematic review on health indicators and policy review on policies for each disease area separately**

1. HIV and Hepatitis B and C

Database(s): **Ovid MEDLINE(R) ALL** 1946 to February 01, 2023
Search Strategy:

| **#** | **Searches** | **Results** |
| --- | --- | --- |
| 1 | exp Human Migration/ | 27701 |
| 2 | exp "Emigrants and Immigrants"/ | 15313 |
| 3 | "Transients and Migrants"/ | 13959 |
| 4 | Refugees/ | 12840 |
| 5 | Refugee Camps/ | 284 |
| 6 | (alien* or asile or asylum* or (border* adj2 cross*) or (countr* adj3 origin*) or diaspora or displace? or displacement* or emigrant* or emigration or expat? or expatriate? or foreigner* or foreign-born* or foreign background* or foreign population* or immigrant* or immigration or migrant* or migration or naturalized citizen* or new* arriv* or newcomer* or new-comer* or nomad* or non-citizen* or nonnative* or non-native* or nonnational or non-national or nonresident or non-resident* or resettlement* or re-settlement* or refugee* or settler* or squatter* or undocumented worker*).ti,ab,kf. | 573213 |
| 7 | or/1-6 | 586984 |
| 8 | exp Middle East/ | 161654 |
| 9 | exp Africa, Northern/ | 40959 |
| 10 | (Abu Dhabi or Ajman or Algeri* or Arab* or Bahrain* or Bahreiin* or Dubai or Egypt* or Emirat* or Fujairah or Gaza* or Golf* or Gulf* or Ifriqiya* or Irak* or Iraq* or Jorda* or Jumhuuriiya* or Koweit* or Kuwait* or Kuwayt* or Leban* or Liban* or Liby* or Lubnan* or Maghr* or Maroc* or Maser* or Masr or Misr or MENA or Middle East* or Morocc* or North* Afric* or Oman* or Palestin* or Qatar* or Saudi* or Sharjah or Soudan* or Sudan* or Syria* or Syuri* or Tunis* or Uman* or Umm Al-Quwain or West Bank or Yemen*).ti,ab,kf. | 317472 |
| 11 | or/8-10 | 436405 |
| 12 | exp Blood-Borne Infections/ or Hepatitis, Viral, Human/ | 428828 |
| 13 | (acquired immunodeficien* syndrome* or AIDS or bloodborne disease* or blood borne disease* or bloodborne infection* or blood borne infection* or Hepatitis B or HBV or Hepatitis C or HCV or HIV or human immunodeficien* virus* or viral hepatitis or viral liver disease*).ti,ab,kf. | 616455 |
| 14 | or/12-13 | 666140 |
| 15 | 7 and 11 and 14 | 721 |
| 16 | (animals not humans).sh. | 5054628 |
| 17 | 15 not 16 | 719 |
| 18 | limit 17 to yr="2000 -Current" | 535 |

2. Malaria and NTDs

Database(s): **Ovid MEDLINE(R) ALL** 1946 to February 24, 2023
Search Strategy:

| **#** | **Searches** | **Results** |
| --- | --- | --- |
| 1 | exp Human Migration/ | 27722 |
| 2 | exp "Emigrants and Immigrants"/ | 15372 |
| 3 | "Transients and Migrants"/ | 14022 |
| 4 | Refugees/ | 12903 |
| 5 | Refugee Camps/ | 288 |
| 6 | (alien* or asile or asylum* or (border* adj2 cross*) or (countr* adj3 origin*) or diaspora or displace? or displacement* or emigrant* or emigration or expat? or expatriate? or foreigner* or foreign-born* or foreign background* or foreign population* or immigrant* or immigration or migrant* or migration or naturalized citizen* or new* arriv* or newcomer* or new-comer* or nomad* or non-citizen* or nonnative* or non-native* or nonnational or non-national or nonresident or non-resident* or resettlement* or re-settlement* or refugee* or settler* or squatter* or undocumented worker*).ti,ab,kf. | 573858 |
| 7 | or/1-6 | 587633 |
| 8 | exp Middle East/ | 159113 |
| 9 | exp Africa, Northern/ | 41057 |
| 10 | (Abu Dhabi or Ajman or Algeri* or Arab* or Bahrain* or Bahreiin* or Dubai or Egypt* or Emirat* or Fujairah or Gaza* or Golf* or Gulf* or Ifriqiya* or Irak* or Iraq* or Jorda* or Jumhuuriiya* or Koweit* or Kuwait* or Kuwayt* or Leban* or Liban* or Liby* or Lubnan* or Maghr* or Maroc* or Maser* or Masr or Misr or MENA or Middle East* or Morocc* or North* Afric* or Oman* or Palestin* or Qatar* or Saudi* or Sharjah or Soudan* or Sudan* or Syria* or Syuri* or Tunis* or Uman* or Umm Al-Quwain or West Bank or Yemen*).ti,ab,kf. | 318043 |
| 11 | or/8-10 | 434382 |
| 12 | Neglected Diseases/ | 2424 |
| 13 | exp Malaria/ or exp Plasmodium/ | 90811 |
| 14 | Buruli Ulcer/ or Mycobacterium ulcerans/ | 1098 |
| 15 | exp Dengue/ or exp Arbovirus Infections/ or exp Flavivirus Infections/ | 52097 |
| 16 | Chikungunya Fever/ | 2723 |
| 17 | Dracunculiasis/ | 907 |
| 18 | exp Echinococcosis/ | 20591 |
| 19 | exp Trematode Infections/ | 37974 |
| 20 | Trypanosomiasis, African/ or Trypanosomiasis/ or Euglenozoa Infections/ | 10112 |
| 21 | exp Leishmaniasis/ | 25341 |
| 22 | exp Leprosy/ or Mycobacterium leprae/ | 24706 |
| 23 | Elephantiasis, Filarial/ or Wuchereria bancrofti/ | 4066 |
| 24 | Mycetoma/ or exp Dermatomycoses/ | 29882 |
| 25 | exp Onchocerciasis/ or exp Filariasis/ or Onchocerca volvulus/ | 16005 |
| 26 | Rabies/ or exp Rhabdoviridae Infections/ | 13177 |
| 27 | Scabies/ or exp Ectoparasitic Infestations/ | 22522 |
| 28 | exp Schistosomiasis/ | 24882 |
| 29 | exp Helminthiasis/ | 134192 |
| 30 | Snake Bites/ | 5405 |
| 31 | exp Taeniasis/ or exp Cestode Infections/ or exp Cysticercosis/ or Neurocysticercosis/ | 33945 |
| 32 | Trachoma/ or exp Conjunctivitis, Bacterial/ | 6181 |
| 33 | Treponemal Infections/ or Yaws/ or Treponema pallidum/ | 5990 |
| 34 | (neglected adj3 disease*).ti,ab,kf. | 8699 |
| 35 | ((arbovirus or echinococcus or euglenozoa or flavivirus or nematomorpha or rhabdoviridae or schistoma or trematode or treponemal) adj3 infection*).ti,ab,kf. | 3845 |
| 36 | (actinomycetoma or african sleeping sickness* or african trypanosomias?s or bacterial conjunctivitis or bancroftian elephantias?s or bancroftian filarias?s or bejel* or bilharsia* or bilharzias?s or black fever or borderline tuberculoid* or breakbone fever or break bone fever or buruli ulcer or chikungunya or coenuri* or coenuros?s or coenurus cerebralis or cysticercos?s or cysticercus cellulosae or deep mycos?s or dengue* or dermatomycos?s or dracuncul* or echinococcos?s or ectoparasitic infestation* or ectoparasitos?s or egyptian ophthalmia or eumycetoma or filarial elephantias?s or filariose or filiariasis or frambesia or guinea worm disease* or hansen* disease* or helminthias?s or hydatid cysts or hydatidos?s or hydrophobia or kalazar or kala azar or katayama fever or leishmani* or leprae or leepre or leprosies or leprosy or lymphatic filarias?s or lyssa or lyssas or madura foot or maduromycosis or malare or malaria or malayi elephantias?s or malayi filarias?s or myceetome or mycetoma or mycobacterium ulcerans or nagana or neurocysticercos?s or neuroschistosomias?s or onchocercias?s or onchocerca volvulus or onchocercose or oriental sore or plasmodium falciparum or paludisme or rabies or raby or river blindness or sarcoptic mange or scabies or schistosomias?s or schistosomal myel* or snakebite* or snake bite* or snake envenoming or taenia* or taenias?s or teeniase or trachoma* or treponema pallidum or treponematos?s or trypanosomiase or trypanosoma or trypanosomiasis or ulceere or ulcer disease* or wuchereria bancrofti or yaw or yaws or zazzabin cizon sauro).ti,ab,kf. | 350022 |
| 37 | or/12-36 | 543044 |
| 38 | 7 and 11 and 37 | 1064 |
| 39 | (animals not humans).sh. | 5062487 |
| 40 | 38 not 39 | 967 |
| 41 | limit 40 to yr="2000 -Current" | 692 |

3. TB

Database(s): **Ovid MEDLINE(R) ALL** 1946 to February 21, 2023
Search Strategy:

| **#** | **Searches** | **Results** |
| --- | --- | --- |
| 1 | exp Human Migration/ | 27721 |
| 2 | exp "Emigrants and Immigrants"/ | 15365 |
| 3 | "Transients and Migrants"/ | 14005 |
| 4 | Refugees/ | 12901 |
| 5 | Refugee Camps/ | 288 |
| 6 | (alien* or asile or asylum* or (border* adj2 cross*) or (countr* adj3 origin*) or diaspora or displace? or displacement* or emigrant* or emigration or expat? or expatriate? or foreigner* or foreign-born* or foreign background* or foreign population* or immigrant* or immigration or migrant* or migration or naturalized citizen* or new* arriv* or newcomer* or new-comer* or nomad* or non-citizen* or nonnative* or non-native* or nonnational or non-national or nonresident or non-resident* or resettlement* or re-settlement* or refugee* or settler* or squatter* or undocumented worker*).ti,ab,kf. | 573334 |
| 7 | or/1-6 | 587109 |
| 8 | exp Middle East/ | 159051 |
| 9 | exp Africa, Northern/ | 41052 |
| 10 | (Abu Dhabi or Ajman or Algeri* or Arab* or Bahrain* or Bahreiin* or Dubai or Egypt* or Emirat* or Fujairah or Gaza* or Golf* or Gulf* or Ifriqiya* or Irak* or Iraq* or Jorda* or Jumhuuriiya* or Koweit* or Kuwait* or Kuwayt* or Leban* or Liban* or Liby* or Lubnan* or Maghr* or Maroc* or Maser* or Masr or Misr or MENA or Middle East* or Morocc* or North* Afric* or Oman* or Palestin* or Qatar* or Saudi* or Sharjah or Soudan* or Sudan* or Syria* or Syuri* or Tunis* or Uman* or Umm Al-Quwain or West Bank or Yemen*).ti,ab,kf. | 317712 |
| 11 | or/8-10 | 433998 |
| 12 | exp Tuberculosis/ | 204795 |
| 13 | (koch* disease or koch's disease or tuberculo?s*).ti,ab,kf. | 248090 |
| 14 | (TB adj3 (active or case* or disease* or infection* or latent)).ti,ab,kf. | 18905 |
| 15 | or/12-14 | 278936 |
| 16 | 7 and 11 and 15 | 472 |
| 17 | (animals not humans).sh. | 5061699 |
| 18 | 16 not 17 | 471 |
| 19 | limit 18 to yr="2000 -Current" | 371 |

4. Vaccine preventable diseases

Database(s): **Ovid MEDLINE(R) ALL** 1946 to February 13, 2023
Search Strategy:

| **#** | **Searches** | **Results** |
| --- | --- | --- |
| 1 | exp Human Migration/ | 27716 |
| 2 | exp "Emigrants and Immigrants"/ | 15329 |
| 3 | "Transients and Migrants"/ | 13990 |
| 4 | Refugees/ | 12879 |
| 5 | Refugee Camps/ | 286 |
| 6 | (alien* or asile or asylum* or (border* adj2 cross*) or (countr* adj3 origin*) or diaspora or displace? or displacement* or emigrant* or emigration or expat? or expatriate? or foreigner* or foreign-born* or foreign background* or foreign population* or immigrant* or immigration or migrant* or migration or naturalized citizen* or new* arriv* or newcomer* or new-comer* or nomad* or non-citizen* or nonnative* or non-native* or nonnational or non-national or nonresident or non-resident* or resettlement* or re-settlement* or refugee* or settler* or squatter* or undocumented worker*).ti,ab,kf. | 572839 |
| 7 | or/1-6 | 586614 |
| 8 | exp Middle East/ | 161966 |
| 9 | exp Africa, Northern/ | 41016 |
| 10 | (Abu Dhabi or Ajman or Algeri* or Arab* or Bahrain* or Bahreiin* or Dubai or Egypt* or Emirat* or Fujairah or Gaza* or Golf* or Gulf* or Ifriqiya* or Irak* or Iraq* or Jorda* or Jumhuuriiya* or Koweit* or Kuwait* or Kuwayt* or Leban* or Liban* or Liby* or Lubnan* or Maghr* or Maroc* or Maser* or Masr or Misr or MENA or Middle East* or Morocc* or North* Afric* or Oman* or Palestin* or Qatar* or Saudi* or Sharjah or Soudan* or Sudan* or Syria* or Syuri* or Tunis* or Uman* or Umm Al-Quwain or West Bank or Yemen*).ti,ab,kf. | 317516 |
| 11 | or/8-10 | 436655 |
| 12 | exp Vaccines/ | 272039 |
| 13 | exp Immunization/ | 205597 |
| 14 | exp Immunization Programs/ | 15956 |
| 15 | (immunostimulation* or immunisation* or immunization* or vaccin* or variolation*).ti,ab,kf. | 461660 |
| 16 | (immunologic* adj2 (sensiti?ation* or stimulation*)).ti,ab,kf. | 603 |
| 17 | Vaccine-Preventable Diseases/ | 225 |
| 18 | Cholera/ | 9238 |
| 19 | COVID-19/ | 213155 |
| 20 | exp Dengue/ | 15622 |
| 21 | Diphtheria/ | 6934 |
| 22 | exp Hepatitis B/ | 64040 |
| 23 | Haemophilus influenzae type b/ or Haemophilus Infections/ | 8717 |
| 24 | exp Papillomaviridae/ | 37470 |
| 25 | Influenza, Human/ | 56964 |
| 26 | Measles/ | 14748 |
| 27 | exp Meningococcal Infections/ | 11761 |
| 28 | Mumps/ | 4847 |
| 29 | Whooping Cough/ | 9095 |
| 30 | exp Pneumococcal Infections/ | 22359 |
| 31 | exp Poliomyelitis/ | 20598 |
| 32 | Rabies/ | 10830 |
| 33 | Rubella/ | 8278 |
| 34 | Rotavirus Infections/ | 8644 |
| 35 | Tetanus/ | 9782 |
| 36 | exp Tuberculosis/ | 204709 |
| 37 | exp Varicella Zoster Virus Infection/ | 19758 |
| 38 | (vaccine preventable adj3 (disease* or illness* or infection*)).ti,ab,kf. | 4284 |
| 39 | (2019 ncov or 2019ncov or breakbone fever or break-bone fever or chickenpox* or chicken pox* or cholera* or cholerae or coqueluche or coronavirus 2 or corona virus disease 2019 or cov2 or cov 2 or covid-19 or covid19 or dengue* or diphtheri* or diphteri* or epidemic parotiti* or german measles or grippe or h1n1 or hav or haemophilus or hbv or hepa or hepatit* or herpesvirus 3 or hepb or hib or hpv or human flu or human papilloma* virus* or hydrophobia or influenza* or koch* disease or lyssa or lyssas or measles or meningit* or meningoco* or mumps or ncov or neisseria meningitidis or neonatal calf diarrhea virus* or new corona virus* or new coronavirus* or novel corona virus* or novel coronavirus* or oreillons or pachymeningiti* or papillomavir* or pertuss* or pfeiffer* bacillus or pneumococcal or pneumoniae or polio* or rabies or rotavirus* or rougeol* or rubella* or rubeol* or sars-cov-2 or sars2 or tdap or tdp or tetani or tetanus* or three day measle* or tuberculos* or varicella or whooping cough*).ti,ab,kf. | 1340987 |
| 40 | or/12-39 | 1744210 |
| 41 | 7 and 11 and 40 | 1725 |
| 42 | (animals not humans).sh. | 5059283 |
| 43 | 41 not 42 | 1682 |
| 44 | limit 43 to yr="2000 -Current" | 1397 |

5. Maternal and neonatal health

Database(s): **Ovid MEDLINE(R) ALL** 1946 to March 23, 2023
Search Strategy:

| **#** | **Searches** | **Results** |
| --- | --- | --- |
| 1 | exp Human Migration/ | 27742 |
| 2 | exp "Emigrants and Immigrants"/ | 15413 |
| 3 | "Transients and Migrants"/ | 14073 |
| 4 | Refugees/ | 12961 |
| 5 | Refugee Camps/ | 287 |
| 6 | (alien* or asile or asylum* or (border* adj2 cross*) or (countr* adj3 origin*) or diaspora or displace? or displacement* or emigrant* or emigration or expat? or expatriate? or foreigner* or foreign-born* or foreign background* or foreign population* or immigrant* or immigration or migrant* or migration or naturalized citizen* or new* arriv* or newcomer* or new-comer* or nomad* or non-citizen* or nonnative* or non-native* or nonnational or non-national or nonresident or non-resident* or resettlement* or re-settlement* or refugee* or settler* or squatter* or undocumented worker*).ti,ab,kf. | 576748 |
| 7 | or/1-6 | 590527 |
| 8 | exp Middle East/ | 159720 |
| 9 | exp Africa, Northern/ | 41145 |
| 10 | (Abu Dhabi or Ajman or Algeri* or Arab* or Bahrain* or Bahreiin* or Dubai or Egypt* or Emirat* or Fujairah or Gaza* or Golf* or Gulf* or Ifriqiya* or Irak* or Iraq* or Jorda* or Jumhuuriiya* or Koweit* or Kuwait* or Kuwayt* or Leban* or Liban* or Liby* or Lubnan* or Maghr* or Maroc* or Maser* or Masr or Misr or MENA or Middle East* or Morocc* or North* Afric* or Oman* or Palestin* or Qatar* or Saudi* or Sharjah or Soudan* or Sudan* or Syria* or Syuri* or Tunis* or Uman* or Umm Al-Quwain or West Bank or Yemen*).ti,ab,kf. | 319881 |
| 11 | or/8-10 | 436540 |
| 12 | Maternal Health/ or Maternal Welfare/ | 8804 |
| 13 | exp Maternal Health Services/ or Maternal-Child Health Centers/ | 58953 |
| 14 | Maternal Mortality/ or Maternal Death/ | 11807 |
| 15 | Hypertension, Pregnancy-Induced/ or Pre-Eclampsia/ | 38475 |
| 16 | Diabetes, Gestational/ | 14923 |
| 17 | Depression, Postpartum/ | 7293 |
| 18 | Postpartum Hemorrhage/ | 8327 |
| 19 | Anemia, Iron-Deficiency/ | 11555 |
| 20 | Pregnancy Complications, Infectious/ or Puerperal Infection/ | 44906 |
| 21 | Pregnancy/ | 978234 |
| 22 | Gravidity/ | 1308 |
| 23 | Pregnancy, High-Risk/ or Pregnancy, Unplanned/ or Pregnancy, Unwanted/ | 9870 |
| 24 | Pregnant Women/ | 13892 |
| 25 | Pregnancy Trimester, Third/ | 15768 |
| 26 | Peripartum Period/ | 1713 |
| 27 | exp Delivery, Obstetric/ or Labor, Obstetric/ or Labor Onset/ or exp Labor Presentation/ or "Trial of Labor"/ or Uterine Contraction/ | 121577 |
| 28 | Birth Setting/ or Home Childbirth/ or Natural Childbirth/ or Term Birth/ | 8431 |
| 29 | Pregnancy Outcome/ or Abortion, Spontaneous/ or Stillbirth/ or Abortion, Incomplete/ or Embryo Loss/ or Abortion, Threatened/ | 84632 |
| 30 | Uterine Cervical Incompetence/ | 1571 |
| 31 | Obstetric Labor Complications/ or Dystocia/ or Premature Birth/ | 41246 |
| 32 | Infant Health/ | 1251 |
| 33 | exp Infant Mortality/ | 31664 |
| 34 | exp Infant, Low Birth Weight/ | 38613 |
| 35 | exp Infant, Premature/ | 64190 |
| 36 | (birth* or child bearing or childbearing or childbirth* or expect* mother* or gestation* or gravidit* or multigravidit* or nulligravidit* or obstetric deliver* or pregnan* or primigravidit*).ti,ab,kf. | 968064 |
| 37 | ((antenatal* or ante natal* or maternal or perinatal* or peri natal* or peripartum or peri partum or postnatal or post natal or postpartum or post partum or puerperium) adj3 (care or clinic* or healthcare or service*)).ti,ab,kf. | 42845 |
| 38 | (abortion* or breech presentation* or dystocia* or embryo death or embryo loss or embryo resorption or fetal presentation* or incompetent cervi* or labor complication* or labor onset* or labor presentation* or labor trial or obstetric labor or prematurity or "trial of labor" or uterine contraction* or uterine cervi* incompetence).ti,ab,kf. | 111733 |
| 39 | ((prematur* or preterm or pre-term) adj3 (baby or babies or child* or deliver* or infant* or neonat* or newborn*)).ti,ab,kf. | 95191 |
| 40 | ((low or small) adj3 (birthweight* or birth weight* or gestational age)).ti,ab,kf. | 51845 |
| 41 | (an?emia* or fetal macrosomia* or gestational diabetes or gestational hypertension or postpartum sepsis or pre-eclampsia or preeclampsia or puerperal infection* or stillbirth*).ti,ab,kf. | 240331 |
| 42 | ((postnatal or post natal or postpartum or post partum) adj2 (depress* or dysphoria* or h?emorrhage*)).ti,ab,kf. | 18564 |
| 43 | ((baby or babies or infant* or maternal* or neonat* or newborn* or peri natal* or perinatal*) adj3 (death* or health* or mortalit*)).ti,ab,kf. | 135076 |
| 44 | or/12-43 | 1658543 |
| 45 | 7 and 11 and 44 | 1881 |
| 46 | (animals not humans).sh. | 5071308 |
| 47 | 45 not 46 | 1843 |
| 48 | limit 47 to yr="2000 -Current" | 1308 |

6. Diabetes

Database(s): **Ovid MEDLINE(R) ALL** 1946 to May 25, 2023
Search Strategy:

| **#** | **Searches** | **Results** |
| --- | --- | --- |
| 1 | exp Human Migration/ | 27778 |
| 2 | exp "Emigrants and Immigrants"/ | 15514 |
| 3 | "Transients and Migrants"/ | 14172 |
| 4 | Refugees/ | 13100 |
| 5 | Refugee Camps/ | 289 |
| 6 | (alien* or asile or asylum* or (border* adj2 cross*) or (countr* adj3 origin*) or diaspora or displace? or displacement* or emigrant* or emigration or expat? or expatriate? or foreigner* or foreign-born* or foreign background* or foreign population* or immigrant* or immigration or migrant* or migration or naturalized citizen* or new* arriv* or newcomer* or new-comer* or nomad* or non-citizen* or nonnative* or non-native* or nonnational or non-national or nonresident or non-resident* or resettlement* or re-settlement* or refugee* or settler* or squatter* or undocumented worker*).ti,ab,kf. | 583255 |
| 7 | or/1-6 | 597041 |
| 8 | exp Middle East/ | 160899 |
| 9 | exp Africa, Northern/ | 41353 |
| 10 | (Abu Dhabi or Ajman or Algeri* or Arab* or Bahrain* or Bahreiin* or Dubai or Egypt* or Emirat* or Fujairah or Gaza* or Golf* or Gulf* or Ifriqiya* or Irak* or Iraq* or Jorda* or Jumhuuriiya* or Koweit* or Kuwait* or Kuwayt* or Leban* or Liban* or Liby* or Lubnan* or Maghr* or Maroc* or Maser* or Masr or Misr or MENA or Middle East* or Morocc* or North* Afric* or Oman* or Palestin* or Qatar* or Saudi* or Sharjah or Soudan* or Sudan* or Syria* or Syuri* or Tunis* or Uman* or Umm Al-Quwain or West Bank or Yemen*).ti,ab,kf. | 323995 |
| 11 | or/8-10 | 441338 |
| 12 | exp Diabetes Mellitus/ | 504402 |
| 13 | exp Insulin Resistance/ | 98730 |
| 14 | (diabet* or insulin* or metabolic syndrome* or cardiometabolic syndrome* or dysmetabolic syndrome* or reaven syndrome*).ti,ab,kf. | 1034882 |
| 15 | or/12-14 | 1092851 |
| 16 | 7 and 11 and 15 | 599 |
| 17 | (animals not humans).sh. | 5090355 |
| 18 | 16 not 17 | 590 |
| 19 | limit 18 to yr="2000 -Current" | 541 |

7. Mental health

Database(s): **Ovid MEDLINE(R) ALL** 1946 to June 08, 2023
Search Strategy:

| **#** | **Searches** | **Results** |
| --- | --- | --- |
| 1 | exp Human Migration/ | 27795 |
| 2 | exp "Emigrants and Immigrants"/ | 15560 |
| 3 | "Transients and Migrants"/ | 14191 |
| 4 | Refugees/ | 13137 |
| 5 | Refugee Camps/ | 291 |
| 6 | (alien* or asile or asylum* or (border* adj2 cross*) or (countr* adj3 origin*) or diaspora or displace? or displacement* or emigrant* or emigration or expat? or expatriate? or foreigner* or foreign-born* or foreign background* or foreign population* or immigrant* or immigration or migrant* or migration or naturalized citizen* or new* arriv* or newcomer* or new-comer* or nomad* or non-citizen* or nonnative* or non-native* or nonnational or non-national or nonresident or non-resident* or resettlement* or re-settlement* or refugee* or settler* or squatter* or undocumented worker*).ti,ab,kf. | 584365 |
| 7 | or/1-6 | 598153 |
| 8 | exp Middle East/ | 161066 |
| 9 | exp Africa, Northern/ | 41399 |
| 10 | (Abu Dhabi or Ajman or Algeri* or Arab* or Bahrain* or Bahreiin* or Dubai or Egypt* or Emirat* or Fujairah or Gaza* or Golf* or Gulf* or Ifriqiya* or Irak* or Iraq* or Jorda* or Jumhuuriiya* or Koweit* or Kuwait* or Kuwayt* or Leban* or Liban* or Liby* or Lubnan* or Maghr* or Maroc* or Maser* or Masr or Misr or MENA or Middle East* or Morocc* or North* Afric* or Oman* or Palestin* or Qatar* or Saudi* or Sharjah or Soudan* or Sudan* or Syria* or Syuri* or Tunis* or Uman* or Umm Al-Quwain or West Bank or Yemen*).ti,ab,kf. | 324777 |
| 11 | or/8-10 | 442219 |
| 12 | Mental Health/ or exp Mental Disorders/ or Depression/ or Anxiety/ or Resilience, Psychological/ | 1608207 |
| 13 | (mental adj2 (disorder* or distress or health or ill-being or illbeing or illness* or ill health or illhealth or instabilit* or symptom* or wellbeing or well-being or wellness)).ti,ab,kf. | 299115 |
| 14 | ((psychiatric or psychologic*) adj3 (disorder* or distress or ill-being or illbeing or ill health or illhealth or illness* or problem* or resilienc* or symptom*)).ti,ab,kf. | 141346 |
| 15 | ((drug* or heroin or inhalant* or marijuana or morphine or opium or substance*) adj3 (abuse* or addict* or dependenc* or habituation*)).ti,ab,kf. | 91981 |
| 16 | (alcoholic intoxication* or alcoholic psychos* or alcoholism or alcohol withdrawal delirium or binge drinking or opiate overdose or substance-induced psychos*).ti,ab,kf. | 39664 |
| 17 | ((alcoholic korsakoff or asperger* or battered child or capgras or creutzfeldt-jakob or female athlete triad or jet lag or kleine-levin or kluver-bucy or munchausen or neonatal abstinence or night eating or restless legs or rumination or substance withdrawal or tourette*) adj3 syndrome*).ti,ab,kf. | 16164 |
| 18 | ((adjustment or affective or alcohol-related or amphetamine-related or attention deficit or auditory perceptual or autistic or autism or bipolar or body dysmorphic or child behavio?r* or (pervasive adj2 development*) or cocaine-related or cognition or combat or communication or conduct or consciousness or excessive somnolence or disruptive behavio?r* or dissociative or "drug use" or eating or elimination or factitious or feeding or gender or hoarding or hyperactivity or impulse control or mood or motor or motor skill* or narcotic-related or neurocognitive or neurodevelopmental or neurotic or obsessive-compulsive or opioid-related or panic or paranoid or paraphilic or personality or phobic or psychotic or sexual or sleep* or sleep wake or somatoform or stress* or substance-related or tic or "tobacco use" or trauma) adj3 disorder*).ti,ab,kf. | 346123 |
| 19 | (agoraphobia or alzheimer disease or amnesia or anorexia nervosa or anxiety or bulimia nervosa or cataplexy or chemotherapy-related cognitive impairment or cognitive dysfunction* or delirium or delusional parasitosis or dementia* or depression* or developmental disabilit* or diabulimia or "diffuse neurofibrillary tangles with calcification" or diurnal enuresis or dyscalculia or dyslexia or dyspareunia or dyssomnia* or emergence delirium or encopresis or enuresis or erectile dysfunction or exhibitionism or firesetting behavior* or food addict* or frontotemporal lobar degeneration or gambling or gender dysphoria* or globus sensation or historical trauma* or huntington disease or hypochondriasis or hysteria* or idiopathic hypersomnia or intellectual disabilit* or kinesiophobia* or learning disabilit* or lewy body disease or masochism or morgellons disease or mutism or narcolepsy or neurasthenia or neurocirculatory asthenia or neurosis or neuroses or night terror* or nocturnal enuresis or nocturnal paroxysmal dystonia or orthorexia nervosa or parasomnia* or pedophilia or phencyclidine abuse* or pica or "pick disease of the brain" or postoperative cognitive complication* or premature ejaculation or primary progressive aphasia or primary progressive nonfluent aphasia* or psychiatric fetishism or psychological sexual dysfunction* or psychological trauma* or pure alexia or "relative energy deficiency in sport" or rem sleep parasomnia* or sadism or schizophrenia or sexual trauma* or sleep bruxism or sleep deprivation* or sleep paralysis or sluggish cognitive tempo or social phobia* or somnambulism or transvestism or trichotillomania or vaginismus or voyeurism or wernicke encephalopathy).ti,ab,kf. | 1001530 |
| 20 | or/12-19 | 2205440 |
| 21 | 7 and 11 and 20 | 2636 |
| 22 | (animals not humans).sh. | 5094204 |
| 23 | 21 not 22 | 2629 |
| 24 | limit 23 to yr="2000 -Current" | 2326 |

**PRISMA study flow diagram**

Studies included in quantitative synthesis meta-analysis)
n =	)

Studies included in qualitative synthesis
n =	)

Full-text articles excluded, with reasons
n =	)

Full-text articles assessed for eligibility
n =	)

Records excluded
n =	)

Records screened
n =	)

Records after duplicates removed
n =	)

Additional records identified through other sources
n =	)

#### Identification

#### Eligibility

#### Included

#### Screening

Records identified through database searching
n =	)
